# Analytical platform to validate microRNA biomarkers for cancer or disease with practically 100% sensitivity and specificity

**DOI:** 10.1101/2024.05.20.24307547

**Authors:** Anastassia Kanavarioti, M. Hassaan Rehman, Salma Qureshi, Aleena Rafiq, Madiha Sultan

## Abstract

We developed a technology for detecting and quantifying trace nucleic acids using a bracketing protocol designed to yield a copy number with approximately ± 20% accuracy across all concentrations. The microRNAs (miRNAs) let-7b, miR-15b, miR-21, miR-375 and miR-141 were measured in serum and urine samples from healthy subjects and patients with breast, prostate, or pancreatic cancer. Detection and quantification were amplification-free and enabled using osmium-tagged probes and MinION, a nanopore array detection device. Combined serum from healthy men (Sigma‒Aldrich #H6914) was used as a reference. Total RNA isolated from biospecimens using commercial kits was used as the miRNA source. The unprecedented ± 20% accuracy led to the conclusion that miRNA copy numbers must be normalized to the same RNA content, which in turn illustrates (i) independence from age, sex, and ethnicity, as well as (ii) equivalence between serum and urine. miR-21, miR-375 and miR-141 copies in cancers were 1.8-fold overexpressed, exhibited zero overlap with healthy samples and had a p value of 1.6×10^-22^, tentatively validating each miRNA as a cancer biomarker. miR-15b was confirmed to be cancer-independent, whereas let-7b appeared to be a cancer biomarker for prostate and breast cancer, but not for pancreatic cancer.

## INTRODUCTION

The incidence of cancer has not declined despite preventive efforts worldwide [1,2]. The cancer incidence rate increases steadily with age and reaches 1 in every 100 people aged ≥ 60 years and older [3]. Most adults undergo an annual or biannual physical medical examination that does not include a multi-cancer test. Specialized tests are available for some prevalent cancer indications, but they are invasive, often painful, and can lead to unacceptable false-positive or false-negative results [4–9]. Rigorous observational studies in Europe failed to determine the effects of mammography screening. Mammography screening results in patients with breast cancer from healthy women and increases the number of mastectomies performed [4,5]. A blood test for prostate-specific antigen (PSA) measures the level of PSA, which is a substance produced by the prostate. The levels of PSA in the blood might be greater in men with prostate cancer, but the PSA test exhibits a 15% false-positive rate, which leads to unnecessary surgical biopsies [6,7]. No single diagnostic test is available for pancreatic cancer. Serum levels of the antigen CA 19-9 higher than 37 U/mL are exploited for the diagnosis of pancreatic cancer in symptomatic patients, but they are not useful as screening markers in asymptomatic individuals because of their low positive predictive value (PPV) [8,9]. A definitive diagnosis requires a series of imaging scans, blood tests, and biopsies, which are typically performed after the appearance of symptoms. Pancreatic cancer is called a “silent” disease because it may cause patients to experience no symptoms until it is too late. Early detection is promising for cancer treatment and for saving lives. Early detection relies heavily on minimally invasive tests using blood samples, urine, saliva, or other biological fluids, so-called liquid biopsies [10,11]. Multiple cancer blood tests using known protein cancer biomarkers [12], circulating tumor DNA shed from tumors [13,14], and DNA methylation profiles [15], are currently being tested in large trials to replace exploratory invasive tissue biopsies and support clinical decisions in symptomatic subjects. Even asymptomatic individuals, especially those aged >60 years, with or without a family history of cancer, are worried about having cancer. A liquid biopsy test to label an asymptomatic individual as cancer-free will ease worries and reduce unnecessary medical procedures. Such a test, like the one proposed herein, will be a valuable addition to regular physical and medical examinations.

**A 2001 seminal publication by Victor Ambros** summarized findings regarding the function of miRNAs [16], a class of small noncoding RNAs with a length of 18 to 25 nucleotides [17,18]. Ambros proposed them to be the “tiny regulators” that control posttranscriptional gene expression, including that related to cell growth, differentiation, development, and apoptosis. miRNAs are abundant in most eukaryotes, and approximately 2500 known miRNAs are common to all humans [18,19]. In 2008, Mitchell et al. illustrated that miRNAs are stable in the blood, which renders them viable biomarkers [20], and empowered more than 150,000 peer-reviewed studies worldwide [21–30]. Several studies have investigated the expression of miR-375 and miR-141 in multiple cancer indications, and many have confirmed that these miRNAs are overexpressed in the serum of cancer patients compared with healthy controls [20,23,26]. A recent review listed 29 medical studies with a total of approximately 7,000 subjects in which elevated miR-21 levels were reported across neoplastic and nonneoplastic diseases [25]. miR-21 may not be a biomarker for a specific disease; however, it is a multi-cancer and multi-disease biomarker. One should be able to determine from a regular medical exam whether his or her miR-21 level is markedly higher than that found in healthy subjects (HL). In addition to miRNAs, no other biomarker has been explored intensely, and found to be involved in cancer onset, progression, metastasis, or survival. Often, the miRNA data from cancer patients overlap with the data from healthy controls, and it is only the median from cancer samples that differs from the median of healthy samples [27,31–35]. For example, droplet digital PCR (ddPCR) data from patients with urological cancers reported copy numbers per μL of plasma for miR-126, miR-141, miR-155, miR-182 and miR-375 in the range of 0-3,000, 0.5-4, 2-40, 1-20 and 1-40, respectively [27]. Notably, a similar significant variation was observed in cancer and healthy samples [27]. This type of data is not atypical for miRNA quantification and has prevented the validation of selected miRNAs as cancer biomarkers.

**The overlap of miRNA data between diseased and healthy subjects,** the quantitative disagreement among studies, and the conjecture that a certain miRNA acts as an oncogene or as a tumor suppressor have been attributed to differences in biospecimen collection methods, study protocols, choice of reference, analytical methods, population variation, disease stage, etc. [31–35]. To improve these statistics, a collective response from an miRNA panel has been proposed. To the best of our knowledge, no miRNA studies have reported zero data overlap between healthy samples and samples with a certain disease. The concentration of miRNAs in blood is in the low femtomolar (fM) range, which is a billion-fold less than the micromolar (μM) range required by typical UV-Vis analytical tools. Current methods for profiling the relative abundance of miRNAs in biological fluids or tissues include small RNA sequencing, reverse transcription-quantitative PCR (RT–PCR), droplet digital PCR (ddPCR), and microarray hybridization [27,31–35]. Although identification works well with these tools, the quantification accuracy and choice of reference have been questioned and may be partially responsible for conflicting conclusions [31–35].

Our earlier work estimated the levels of miR-21, miR-375, and miR-141 in cancer to be 2 to 3-fold higher compared to those in healthy controls. This estimate agrees with the 1.8-fold overexpression observed in a prostate cancer study [23], and the 1.7-fold overexpression of miR-21 observed in a lung cancer study [22]. The data described below were in excellent agreement with those reported in previous studies. A 2-fold overexpression is a small effect that is difficult to measure accurately using amplification-based techniques. This may explain the observed disagreement between studies and the overlap of data between cancer and healthy samples. A tentative cancer screening assay for asymptomatic subjects must be robust, reliable, and accurate and should be preceded by studies with zero data overlap between cancer and healthy samples.

To achieve zero data overlap, an analytical assay for biomarker overexpression must exhibit a lower limit in cancer samples that is equal to or higher than the upper limit observed in healthy controls. Similarly, an analytical assay testing biomarker underexpression must exhibit a higher limit in cancer samples that is equal to or lower than the lower limit in healthy controls. Tables 1A and 1B illustrate the interplay between assay accuracy and biomarker/miRNA overexpression or underexpression. Notably, over- or under-expression of the specific biomarker did not change the correlation between x-fold expression and assay accuracy (see identical results in the last columns of Tables 1A and 1B). For example, if an miRNA quantification assay is associated with a ± 30% accuracy, then a disease associated with less than 1.85-fold miRNA overexpression will yield overlapping data between healthy and diseased samples, and this miRNA will not be validated as a biomarker. Alternatively, if the same assay investigates a disease associated with more than 1.85-fold miRNA overexpression, the data will exhibit zero overlap between healthy and diseased individuals. These correlations presume that the only significant parameter for miRNA levels is the presence or absence of a disease. It was also assumed that assay accuracy remained constant within the range of the tested measurements. Other parameters, like age, gender, race, etc., if turn out to be significant, will need to be addressed separately by limiting the tested population. Typically, analytical assays require extensive real-life sample testing to determine assay accuracy, which delays development and implementation. An analytical tool with protocol-defined accuracy, constant across all relevant concentrations, such as that described below, will streamline testing, and enable miRNA validation.

**Table 1A.** Correlation of measurement’s accuracy and miRNA x-fold overexpression to achieve zero overlap between healthy and diseased samples (1). (1) To understand the examples shown in the table, let us consider the calculation for ± 20% accuracy. The average normalized control was 1.0, with a lower limit (0.8) and an upper limit (1.2). Note that the ratio of the upper limit to the lower limit is 1.2/0.8 = 1.5. A zero overlap of data requires that the lower limit of disease measurement be equal to or greater than 1.2. For a normalized overexpressed disease, the same 20% accuracy yields a factor of 1.5, with a lower limit of 1.2 and an upper limit of 1.8. The other entries in this table are calculated in a similar manner.

| Accuracy of measurement (+/-) | Normalized control, range | Average control | Normalized disease overexpressed, range | Average disease | miRNA in disease/control, x-fold |
| --- | --- | --- | --- | --- | --- |
| 15% | 0.85 to 1.15 | 1.0 | 1.15 to 1.55 | 1.35 | > 1.35 |
| 20% | 0.8 to 1.2 | 1.0 | 1.2 to 1.8 | 1.5 | > 1.5 |
| 30% | 0.7 to 1.3 | 1.0 | 1.3 to 2.4 | 1.85 | > 1.85 |
| 40% | 0.6 to 1.4 | 1.0 | 1.4 to 3.2 | 2.3 | > 2.3 |
| 50% | 0.5 to 1.5 | 1.0 | 1.5 to 4.5 | 3.0 | > 3.0 |
| 75% | 0.25 to 1.75 | 1.0 | 1.75 to 12.25 | 7.0 | > 7.0 |

**Table 1B.** Correlation of measurement’s accuracy and miRNA x-fold under-expression to achieve zero overlap between healthy and diseased samples (1). (1) Small numerical differences between the last columns in Tables 1A and 1B are due to rounding up of the calculations. For calculations see footnote under Table 1A, whereas the lower limit of the normalized control range serves as the upper limit of the normalized disease range, which is under-expressed.

| Accuracy of measurement (+/-) | Normalized control, range | Average control | Normalized disease Underexpressed, range | Average disease | miRNA in control/disease, x-fold |
| --- | --- | --- | --- | --- | --- |
| 15% | 0.85 to 1.15 | 1.0 | 0.63 to 0.85 | 0.74 | > 1.35 |
| 20% | 0.8 to 1.2 | 1.0 | 0.54 to 0.8 | 0.67 | > 1.5 |
| 30% | 0.7 to 1.3 | 1.0 | 0.4 to 0.7 | 0.55 | > 1.8 |
| 40% | 0.6 to 1.4 | 1.0 | 0.3 to 0.6 | 0.45 | > 2.2 |
| 50% | 0.5 to 1.5 | 1.0 | 0.18 to 0.5 | 0.34 | > 2.9 |
| 75% | 0.25 to 1.75 | 1.0 | 0.036 to 0.25 | 0.14 | > 7.1 |

The diverging literature reports and correlations in Tables 1A and 1B highlight the need for a novel analytical tool. We opted for an amplification-free assay, a nanopore-array detector suitable for single-stranded (ss) nucleic acid trace measurements, and a bracketing protocol designed to deliver miRNA copies with approximately ± 20% accuracy across all concentrations. The proof of principle of this concept was reported in 2020 [36]. A recent study highlighted the implementation of this novel technique for quantifying miRNAs in serum and estimated 2- to 3-fold overexpression of known miRNA cancer biomarkers [37]. Here, we confirmed earlier serum results and, using let-7b as an example, detailed an experimental strategy for validating miRNAs as cancer biomarkers. Strikingly, miRNA measurements using serum and urine samples demonstrated equivalence, which advocates the replacement of blood drawn for urine collection. The technology implemented here is well positioned to overcome inconsistencies in the miRNA field and revolutionize the current thinking in validating miRNA biomarkers.

**Nanopores have shown promise for trace measurements,** and experimental platforms have been successfully used to quantify miRNAs [38–46]. The MinION device from Oxford Nanopore Technologies (ONT) is the only commercially available nanopore array detection device used in conjunction with a consumable flow cell with 2048 embedded nanopores. These proprietary protein nanopores permit the translocation of ss and prevent the translocation of double-stranded (ds) nucleic acids, as shown by ONT and us. The platform includes 512 independent detection channels, with one detection channel per four nanopores, and is promoted for sequencing long ss DNA/RNA. Sequencing combined with DNA-barcoded probes has been used for the detection of biomarkers, including miRNAs [47,48]. As an alternative, rolling circle reverse transcription was used to sequence miRNAs using MinION [49]. However, the lowest detection level of these techniques is in the picomolar range, which is 100- to 1000-fold higher than the miRNA levels in biological fluids, rendering these techniques unsuitable for miRNA quantification in liquid biopsies.

**The MinION software, MINKNOW,** reports the raw data, that is, the ion current (i), as a function of time (t) and can be exploited for ion conductance (sensing) experiments, as described by us [36,37,46] and others [50]. Our earlier study showed that selective osmium tagging of an oligo yields a chemically stable probe that hybridizes efficiently with complementary DNA, RNA, or miRNA targets. These probes traverse size-appropriate proteins [45] and solid-state nanopores [44], including nanopores on the MinION platform [36,37,46]. Owing to the bulkiness of the osmium tags, the translocation of the probe was markedly slower than that of the intact RNA/DNA. Whereas other nanopore platforms can detect and report all translocations, MinION selectively detects our probes over intact nucleic acids. This is because of the relatively slow data acquisition rate at three data points/ms, which quantitatively detects our optimized probes but misses most intact nucleic acids. In the absence of the target, the probe is free, traverses the nanopores and is detected owing to an increase in the reported events over the background noise (see Figure 1, scheme in the middle). Hybridization between the target and the probe yields a hybrid that is too large to traverse the nanopore, resulting in no probe detection or silencing. Target quantification was based on 1:1 hybridization and a known probe concentration. The number of miRNA copies was determined by bracketing, that is, from the average of two experiments, one that yielded probe detection and another that yielded no probe detection, or silencing [36,37]. To obtain the desired ± 20% accuracy, the two experiments must differ by approximately 67% in either the probe or RNA (see discussion). Typically, more than two experiments, were conducted to confirm the miRNA copy number determination. Therefore, the assay is currently low throughput but delivers data with a protocol defined by ± 20% accuracy.

**Figure 1:**
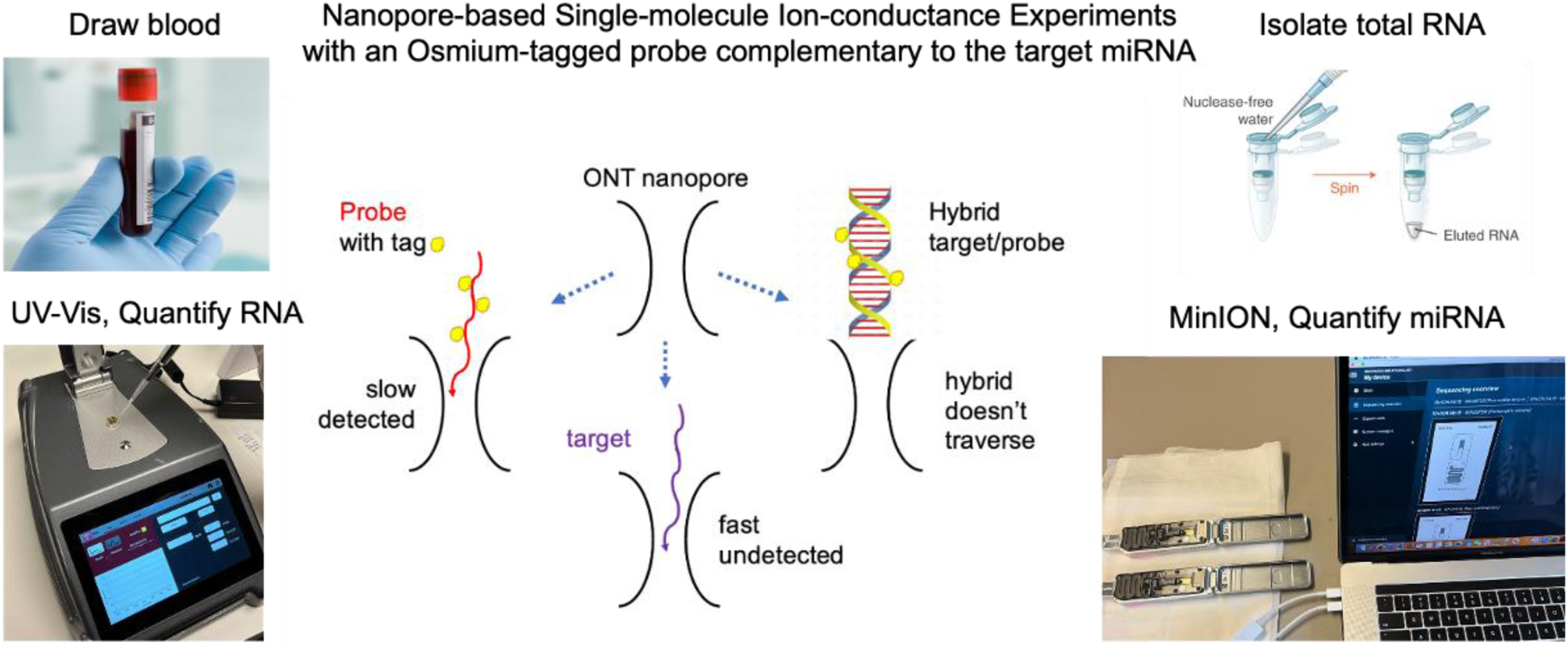
Graphical abstract of all the processes involved in the miRNA measurement using the MinION platform. From left to right: (i) collection of the biospecimen, blood or urine; (ii) isolation of total RNA using a commercial kit; (iii) measurement of total RNA in the isolate using a DeNovix DS-11 spectrophotometer; and (iv) mixing of an aliquot from the RNA isolate with an aliquot of the probe complementary to the target miRNA, adding ONT buffer and conducting a MinION ion-conductance experiment (two experiments running simultaneously, shown here). The experiment measures the ion current (*I*) in picoAmperes (pA) as a function of time (t) in milliseconds (ms). In practice, *I* is constant at *I_o,_*which is the open nanopore ion current (*I*_o_). When a single molecule traverses the nanopore, *I_o_* is reduced to a new value, *I_r_*, because the molecule occupies the space that would have been occupied by the electrolyte that produces *I_o_*. Ion current reduction (dip in this platform) lasts for a time, τ; both *I_r_* and τ depend on the molecular characteristics. The data were stored automatically as a fast5 file, which was subsequently analyzed by *OsBp_detect* (our software, see below). The analysis determines whether the free probe is in excess and detected (left on the scheme above) or if the probe is not detected because it is hybridized with the target (right on the scheme above). Notably, RNAs, including the target miRNA, traverse much faster than the probes, and they are not detected (bottom on the scheme above) due to the relatively slow acquisition rate of this platform.

## RESULTS AND DISCUSSION

### ONT provides protocols for sequencing long RNAs/DNAs but not for single molecule, ion conductance (sensing)

The ONT software (MINKNOW) recorded the raw data from the ion conductance experiments, which were subsequently analyzed using a publicly available algorithm, *OsBp-detect,* developed specifically for this application [51]. While most studies report relative miRNA abundance, our technology measures miRNA copies in aliquots used for the nanopore experiments. Mitchell et al. reported miR-15b at ∼10,000 and miR-16 at ∼110,000 copies per 1 μL of plasma from three individuals [20]. We recently reported that miR-15b = 8,855 and miR-16 = 105,125 copies per 1 μL of H6914 combined serum [37 and Table 2]. This consensus was the first demonstration of a MinION-based sensing assay. This agreement was partially fortuitous because the total RNA from the serum exhibited rather small sample-to-sample variation [52], rendering the H6914 RNA content comparable to the plasma RNA content of the individuals tested by Mitchell et al. [20]. Over the last three years, we purchased four different lots of H6914, isolated total RNA (∼16ng/μL) from each and found reproducible copy numbers for six miRNAs (Table 2). This consistency is remarkable considering that (i) total RNA was isolated using different lots of the Monarch RNA isolation kit from New England Biolabs; (ii) nanopore measurements were conducted by different analysts; (iii) chemically distinct probes were used; (iv) different experimental protocols were used; (v) different versions of MinION flow cells (R9 or R10) were used; and (vi) different versions of MINKNOW software were used.

**For privacy reasons and to circumvent a blood draw, urine was explored as the miRNA source,** as miRNAs have been found to be relatively stable in urine [53]. To directly assess whether miRNAs exhibit comparable copy numbers in serum and urine, we initially purchased a set of 12 samples, matched serum and urine samples from the same donor, and two donors each for breast, prostate, and pancreatic cancer. Preliminary data support the hypothesis that urine can replace blood. However, the 1 mL urine volume afforded 0.05 mL of total RNA at concentration of approximately 7 ng/μL, which is at the lower limit of our technology, and was not sufficient to run the number of experiments required to reach solid conclusions. We did not pursue this type of matching serum and urine study further because of the striking equivalence between miRNA copy numbers determined from H6914 serum and urine samples from healthy subjects (see later). Urine may contain up to 50-fold less RNA than the serum. While 0.2 mL of serum provides 0.1 mL of isolated total RNA sufficient for multiple miRNA determinations, a much larger urine volume is necessary. A recently developed slurry kit from Norgen Biotek enabled the isolation of a 0.05 mL sample of total RNA from 5 to 10 mL of urine. miR-16 was found to be 12-fold more abundant in serum than miR-15b [20,37]. Attempts to measure miR-16 in urine failed, suggesting that miR-16 may be under-expressed in urine compared to serum. The other five miRNAs were measured in both the serum and urine samples (see Tables 2-4).

**Table 2.**
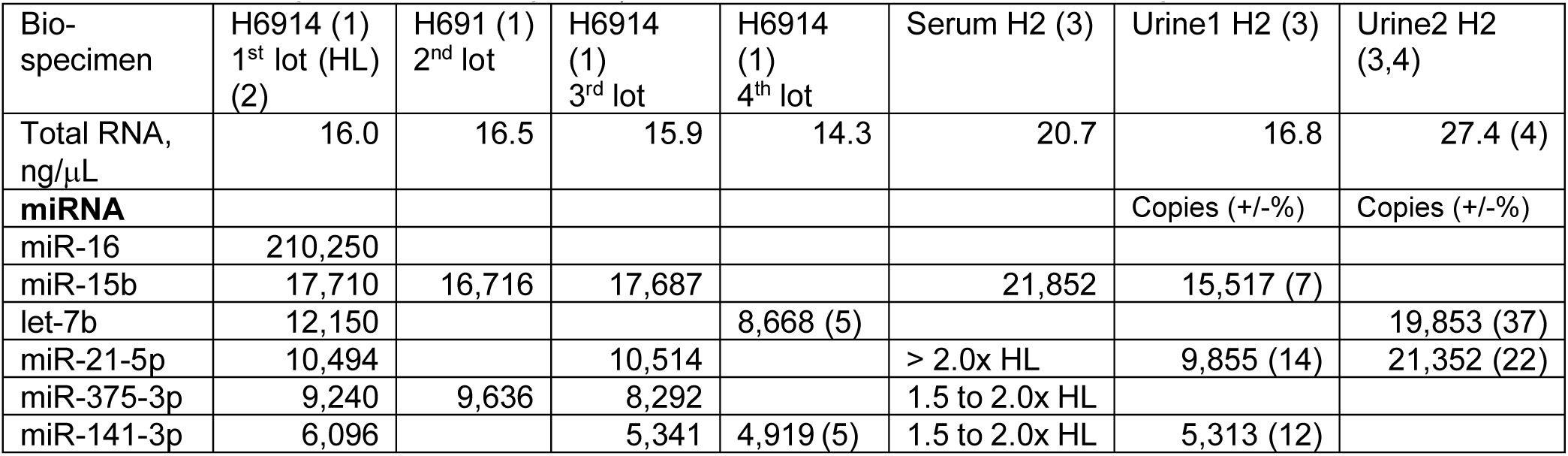
miRNA copies measured per 1 μL of total RNA isolated from a biospecimen, serum or urine. (1) H6914 was the combined serum of healthy men and was purchased from Sigma‒Aldrich. miRNA copies measured from the first and second lots were reported earlier per 1μL of serum [37] and are listed here per 1μL of total RNA which equals 2μL of serum. The accuracy values not listed here are all less than +/-20%. (2) HL stands for Healthy Level (H6914 1^st^ lot is the control/reference in this study). (3) H2 is a healthy woman, in the age group 71-75, tested multiple times over a period of 3 years (see Tables 3 and 4); only the serum HW data were reported earlier [37]. H2 (female) miRNA copies (six measurements for five miRNAs) normalized to the RNA content of the H6914 1^st^ lot are listed in Table 4 for both urine samples and illustrate the match with H6914. (4) The original Urine2 sample contained RNA 82.3 ng/μL, which was diluted 1/3 with water before mixing with the probe. The number of miR-21 copies normalized to the RNA content in the H6914 1^st^ lot was 21,352×16/27.4=12,468. The number of copies of H2-related miR-21 normalized to the number of copies of miR-21 in the H6914 1^st^ lot was 12,468/10,494=1.19 (Table 3, 5^th^ row, in the Healthy Urinary section). (5) Normalized to the 1^st^ lot of H6914 yielded let-7b=9,698 (HL=0.80) from 8,668×16/14.3 (9,698/12,150), and miR-141=5,503 (HL=0.90) from 4,919×16.0/14.3 (5,503/6,096), both within experimental error comparable to the other lots.

**Table 3:** MinION experiments targeting let-7b in healthy and cancer samples to illustrate validation strategy of let-7b as prostate and breast cancer biomarker, but not for pancreatic cancer. (1) The logistics of the subjects are listed in Table 5. B at the sample ID represents the 2^nd^ total RNA isolation. Samples with RNA concentrations > 35 ng/μL were diluted with Ambion water as shown. (2) PAN, PRO, and BRE represent pancreatic, prostate and breast cancer, respectively. (3) Aliquots of total RNA and probe used for the ion conductance nanopore experiment. Probe for let-7b at 30 fM tagged with an average of 5.5 OsBp moieties (Table 6 in Methods). (4) Normalization to the same RNA content (16ng/μL H6914, 1^st^ lot). Probe copies P were calculated from P=600 x probe (μL) x 30 (fM) (5) Dividing the content normalized probe copies by the let-7b copy number (12,150 copies, H6914, 1^st^ lot) to obtain x-fold healthy level (HL); see also under (8). (6) The assignment of an experiment as detection or silencing is based on the R-factor, which is the ratio of the event counts of late (Ir/Io)max to early (Ir/Io)max (see Figures and Methods for discussion and [37]). One test using this technology comprised of three nanopore experiments (45 min each at -180 mV). The first experiment is the baseline or control experiment and is typically an experiment with a buffer only. The mixture sample (RNA+probe) was then loaded onto the flow cell and run twice on the same flow cell under the above conditions. The results of these three experiments are compared, and R for each is determined in the order of control, 1^st^ run, and 2^nd^ run. (7) SIL and DET stand for Silencing and Detection, respectively. The R values of the 1^st^ and 2^nd^ runs were compared to the control and if, at least, one of them decreased, then the experimental result was “detection”. If at least one of them increases, then the experimental result is “silencing”. If one of the runs is an increase and the other a decrease, the experiment is considered currently inconclusive (Note 1), even though preliminary evidence suggests that this experiment directly yields the miRNA copy number. R decreasing or increasing must be statistically significant. (8) The two experiments with an asterisk are used to determine the let-7b copy number, and the other entries are confirmatory. For a specific experiment with an x μL probe and y μL of sample RNA, probe molecules P = x μL (probe concentration in fM) × 600. If the experiment involved detection, then P > target miRNA molecules within the y μL aliquot. If the experiment involved silencing, then P < target miRNA molecules in y μL. miRNA copies per 1 μL of isolated RNA sample < or > P/y, depending on the experimental outcome.

| subject (1) | bio-specimen | condition (2) | isolated total RNA (ng/μL) | RNA (μL) (3) | probe. let-7b (μL) (3) | probe copies per 1 μL of RNA | Normalized probe copies (4) | Normalized probe copies/ 12,150 (5) | Ratio of late over early (lr/lo)max (6) | experimental result (7) | let-7b / let-7b HL (8) | x-fold HL (+/-) |
| --- | --- | --- | --- | --- | --- | --- | --- | --- | --- | --- | --- | --- |
| H1 | urine | healthy | 18.8 | 9.5 | 6.0 | 11,368 | 9,675 | 0.80 | R=3.5, 4.8, 9.0 (increase x2) | SIL | > 0.80 HL * |  |
| " | " | " | " | 6.3 | 6.0 | 17,143 | 14,590 | 1.20 | R=2.4, 1.2, 1.2 (decrease x2) | DET | < 1.20 HL * | 1.00 HL (0.20) |
| H2 1/3 dilution | " | " | 27.4 | 6.0 | 6.0 | 18,000 | 10,499 | 0.86 | R=2.1, 3.9, 1.1 (increase x1, decrease x1) | Note 1 |  |  |
| " | " | " | " | 4.0 | 6.0 | 27,000 | 15,749 | 1.3 | R=6.7, 4.3, 1.9 (decrease x2) | DET | < 1.30 HL * |  |
| " | " | " | " | 4.0 | 6.0 | 27,000 | 15,749 | 1.30 | R=1.6, 1.5, 1.1 (decrease x1) | DET | < 1.30 HL * |  |
| " | " | " | " | 8.5 | 6.0 | 12,706 | 7,411 | 0.61 | R=4.1, 3.7, 5.4 (increase x1) | SIL | > 0.61 HL * | 1.07 HL (0.37) |
| H6914 4th lot | serum men | " | 14.3 | 10.5 | 5.0 | 8,571 | 9,590 | 0.79 | R=2.2, 1.5, 2.3 (decrease x1) | DET | < 0.79 HL * |  |
| " | " | " | " | 6.8 | 5.0 | 13,235 | 14,809 | 1.22 | R=3.5, 3.8, 2.4 (decrease x1) | DET | < 1.22 HL |  |
| " | " | " | " | 13.0 | 5.0 | 6,923 | 7,746 | 0.64 | R=1.0, 1.5, 1.0 (increase x1) | SIL | > 0.64 HL * | 0.72 HL (0.08) |
| SR16-690 | serum | PAN cancer | 16.4 | 9.0 | 5.0 | 10,000 | 9,756 | 0.80 | R=1.2, 1.7, 1.2 (increase x1) | SIL | > 0.80 HL * |  |
| " | " | " | " | 6.0 | 5.0 | 15,000 | 14,634 | 1.20 | R=5.3, 2.0, 1.9 (decrease x2) | DET | < 1.20 HL * |  |
| " B | " | " | 14.3 | 6.5 | 5.0 | 13,846 | 15,492 | 1.28 | R=3.6, 1.9, 1.6 (decrease x2) | DET | < 1.28 HL |  |
| " B | " | " | " | 13.0 | 5.0 | 6,923 | 7,746 | 0.64 | R=1.0, 1.6, 0.8 (increase x1) | SIL | > 0.64 HL | 1.00 HL (0.20) |
| SR17-248 B | " | " | 21.0 | 4.5 | 5.0 | 20,000 | 15,238 | 1.25 | R=1.6, 0.9, 1.6 (decrease x1) | DET | < 1.25 HL |  |
| " | " | " | " | 7.5 | 5.0 | 12,000 | 9,143 | 0.75 | R=1.1, 0.6, 0.7 (decrease x2) | DET | < 0.75 HL * |  |
| " | " | " | " | 9.5 | 5.0 | 9,474 | 7,218 | 0.59 | R=1.2, 2.3, 2.2 (increase x2) | SIL | > 0.59 HL * | 0.67 HL (0.08) |
| SR23-6022 | urine | PRO cancer | 14.5 | 4.0 | 5.0 | 22,500 | 24,828 | 2.04 | R=3.7, 3.0, 1.0 (decrease x2) | DET | < 2.04 HL * |  |
| " | " | " | " | 5.0 | 4.0 | 14,400 | 15,890 | 1.31 | severely reduced events | SIL | > 1.31 HL |  |
| " | " | " | " | 8.0 | 4.0 | 9,000 | 9,931 | 0.82 | severely reduced events | SIL | > 0.82 HL |  |
| " | " | " | " | 4.0 | 4.0 | 18,000 | 19,862 | 1.63 | severely reduced events | SIL | > 1.63 HL * | 1.82 HL (0.20) |
| " | " | " | " | 5.3 | 5.0 | 16,981 | 18,738 | 1.54 | R=1.6, 1.8, 1.3 (comparable) | Note 1 |  |  |
| SR23-6028 | " | " | 12.4 | 10.0 | 4.0 | 7,200 | 9,290 | 0.76 | R=0.7, 0.9, 1.7 (increase x1) | SIL | > 0.76 HL |  |
| " | " | " | " | 6.0 | 4.0 | 12,000 | 15,484 | 1.27 | R=1.9, 1.5, 2.7 (increase x1) | SIL | > 1.27 HL * |  |
| " | " | " | " | 4.7 | 4.0 | 15,319 | 19,767 | 1.63 | R=2.7, 1.7, 2.0 (decrease x2) | DET | < 1.63 HL * | 1.45 HL (0.18) |
| SR23-6016 1/4 dilution | " | BRE cancer | 43.7 | 4.0 | 6.0 | 27,000 | 9,886 | 0.81 | R=1.5, 2.9, 2.3 (increase x2) | SIL | > 0.81 HL |  |
| " | " | " | " | 2.7 | 6.0 | 40,000 | 14,645 | 1.21 | R=2.0, 2.3, 2.4 (comparable) | Note 1 |  |  |
| SR23-6016 1/8 dilution | " | " | 21.8 | 4.0 | 7.5 | 33,750 | 24,771 | 2.04 | R=6.6, 2.2, 3.6 (decrease x2) | DET | < 2.04 HL * |  |
| " | " | " | " | 4.0 | 6.0 | 27,000 | 19,817 | 1.63 | R=1.7, -, 3.2 (increase x1) | SIL | > 1.63 HL * | 1.77 HL (0.28) |
| " | " | " | " | 5.5 | 7.5 | 24,545 | 18,015 | 1.48 | R=1.1, 1.9, 3.2 (increase x2) | SIL | > 1.48 HL |  |
| 101499 | serum | " | 17.1 | 5.5 | 6.0 | 19,636 | 18,373 | 1.51 | R=3.8, 3.9, 4.3 (increase x1) | SIL | > 1.51 HL * |  |
| " | " | " | " | 4.0 | 6.0 | 27,000 | 25,263 | 2.08 | R=1.5, 0.8, 0.9 (decrease x2) | DET | < 2.08 HL * | 1.80 HL (0.28) |

**Table 4:** miRNA copies (HL units, Figure 2) from experiments normalized as described in Table 3. (1) The logistics of the subjects are listed in Table 5. Logistics for H2 sample are in the footnote of Table 2. (2) Multiple entries of RNA isolated from healthy samples corresponded to timely separate collections. When the amount of isolated RNA exceeded 35 ng/μL, it was diluted with Ambion water before mixing with the probe. (3) Cancer serum samples (second, third, and fourth row entries) were also used in an earlier study [37]; however, only miR-15b was measured earlier. The same RNA isolate was used for the additional miRNAs measured here. As described in Table 3, measured probe copies were normalized to the same RNA content and then divided by the corresponding miRNA copy number using as reference H6914 1^st^ lot (Table 2). This yielded HL levels from which copy number is obtained and listed in columns 4 through 8 as x-fold HL. The last column reports the results of a single experiment in which two miRNAs were simultaneously targeted (see Discussion section). Experiments yielding silencing or detection, not followed by the corresponding test to determine miRNA copy number, are not included here, but were confirmatory.

| ID (1) | indication | Isolated total RNA, ng/μL (2) | miRNA targets in HL units (3) |  |  |  |  |
| --- | --- | --- | --- | --- | --- | --- | --- |
|  |  |  | miR-15b | miR-21 | miR-375 | miR-141 | miR-375 + miR-141 |
| Cancer serum |  |  |  |  |  |  |  |
| CAN7 | breast | 6.9 | 0.79 | 1.60 |  |  |  |
| CAN9 | “ | 9.1 | 0.89 | 1.79 |  | 1.80 |  |
| CAN4 | prostate | 12.0 | 0.88 | 1.79 |  | 1.81 |  |
| CAN6 | “ | 8.0 | 0.90 | 1.87 |  | 1.84 |  |
| SR16-690 | pancreatic | 16.4 | 1.01 | 1.63 |  |  |  |
|  |  |  |  | 1.88 |  |  |  |
| SR17-248 | pancreatic | 14.4 | 1.00 |  |  | 1.88 |  |
| Cancer urine |  |  |  |  |  |  |  |
| SR23 6016 | breast | 174.6 | 1.34 | 1.75 |  | 1.76 | 1.69 |
| SR23 6017 | “ | 88.8 | 1.12 | 2.13 |  | 1.73 | 1.66 |
|  |  |  |  |  |  | 2.20 |  |
| SR23 6018 | breast | 16.1 |  |  |  | 1.72 |  |
|  |  |  |  |  |  | 2.25 |  |
| SR23 6022 | prostate | 15.3 |  |  | 1.81 | 1.80 |  |
| SR23 6028 | “ | 13.3 |  |  | 1.81 | 1.82 |  |
| SR23 6023 | “ | 18.5 |  |  | 1.82 | 1.63 |  |
|  |  |  |  |  |  | 2.00 |  |
| SR23 6033 | pancreatic | 13.4 |  |  | 1.82 | 1.82 |  |

| Healthy urine |  |  |  |  |  |  |  |
| --- | --- | --- | --- | --- | --- | --- | --- |
|  |  | 7.5, 22.9 | 0.97 |  | 1.04 |  |  |
|  |  | 21.7 |  |  | 0.97 |  |  |
|  |  | 9.5 | 1.00 |  |  |  |  |
| H2 |  | 16.8, 82.3 | 0.84 | 0.90 |  | 0.83 | 0.84 |
|  |  |  |  | 1.19 |  |  |  |
|  |  | 8.7 |  | 1.01 |  |  |  |
|  |  | 16.5 |  |  |  |  | 0.89 |
|  |  | 12.2 | 1.00 |  |  |  |  |
|  |  | 57.4 | 0.98 |  |  |  | 0.80 |
|  |  | 163.0 | 0.84 |  |  | 1.02 |  |
|  |  | 11.7 |  |  |  |  | 0.96 |
|  |  | 25.6 |  | 1.03 |  |  | 0.87 |
|  |  | 14.5 |  |  |  |  | 0.92 |
|  |  | 16.4 |  | 1.17 |  |  | 1.16 |
|  |  | 13.8 | 0.79 | 1.17 |  |  | 1.06 |
|  |  | 12.4, 15.7 | 1.02 | 1.42 |  |  | 1.30 |
|  |  | 14.7 |  | 1.30 |  |  | 1.31 |

A groundbreaking discovery was made when miRNA copies in the serum and urine (Urine1) of a healthy woman (H2) were found to be equivalent to the corresponding miRNA copies in the H6914 serum (Table 1). A second urine sample from H2 (Urine2) confirmed these findings after normalization to the H6914 RNA content (see footnote in Table 1). Notably, the three samples from H2 were collected months apart and had distinct RNA content. To the best of our knowledge this is the first demonstration of miRNA copy number equivalence between healthy serum (men, combined) and healthy urine (woman) collected from different sources.

**The samples tested in this study** included H6914 (3^rd^ and 4^th^ lots), serum and urine samples from cancer patients, and urine samples from healthy subjects. The miRNA/urine study was approved by the Advarra IRB (see Methods). The instructions for urine collection were identical for both healthy and diseased women and men. No formal follow-up was planned in the healthy group. Breast, prostate and pancreatic cancer samples were purchased from two blood banks, Discovery Life Sciences and Tissue for Research, with subject requirements for early-stage diagnosis (I or II) and before treatment, because miRNA levels may be influenced by disease stage and therapy. The selection of an early disease stage ahead of treatment is consistent with our objective of providing a validation strategy for miRNA biomarkers and developing a cancer-screening test for asymptomatic individuals. The cancer samples were selected to be as inclusive as possible with respect to age, sex, and ethnicity (see Table 5 in Methods). Healthy urine samples were obtained from subjects who varied in sex and ethnicity and ranged in age from 30 to 75 years.

**Table 5:** Demographics of cancer patients whose samples are listed in Tables 3 and 4.

| biobank | ID | Age group | gender | cancer | T stage | N Stage | specimen |
| --- | --- | --- | --- | --- | --- | --- | --- |
| Tissue for Research, UK | 101499 | 56-60 | F | breast | - | - | serum matched |
|  | SR23 6016 | 51-55 | F | " | pT1b | pN0 | urine |
|  | SR23 6017 | 66-70 | F | " | pT1b | " | " |
|  | SR23 6018 | 51-55 | F | " | pT1a | " | " |
|  | SR23 6022 | 71-75 | M | prostate | pT2 | " | " |
|  | SR23 6023 | 66-70 | M | " | " | " | " |
|  | SR23 6028 | 51-55 | M | " | " | " | " |
|  | SR23 6033 | 66-70 | F | pancreatic | " | " | " |
|  | SR16 690 | 51-55 | M | " | pT2 | " | serum |
|  | SR17 248 | 51-55 | M | " | pT1 | " | " |
| Discovery Life Sciences, US | CAN4 | 66-70 | M | prostate | newly diagnosed, pretreatment |  | serum |
|  | CAN6 | 56-60 | M | " |  |  | " |
|  | CAN7 | 51-55 | F | breast |  |  | " |
|  | CAN9 | 56-60 | F | " |  |  | " |

### Validation strategy illustrated by targeting let-7b in three cancer indications

For simplicity only a small number of samples were tested here, even though validation should include at least 10 samples each from healthy subjects and subjects diagnosed with a certain disease ahead of treatment, as mentioned earlier. The first three samples in Table 3 were from healthy subjects, while the 4^th^ and 5^th^ samples were serum samples from patients with pancreatic cancer. Within the accuracy of this technology, these five samples provided statistically indistinguishable let-7b copy numbers. To simplify the comparison, copy numbers were normalized to the H6914 1^st^ lot RNA content of 16.0 ng/μL (reference), and then divided by the let-7b copy number (12,150) from this reference sample to give the HL number posted in the last column of Table 3. These five numbers (1.00, 1.07, 0.72, 1.00 and 0.67) give an average 0.89 HL with RSD=0.21. The tentative conclusion is that let-7b is not a pancreatic cancer biomarker and will be useful in discriminating pancreatic cancer from breast and prostate cancer. This finding should be confirmed using additional samples. The 6^th^ and 7^th^ samples are urine samples from patients diagnosed with prostate cancer with let-7b copy number at 1.82 and 1.45 HL (HL from H6914 1^st^ lot). Compared to this study’s control at 0.89 HL, the 6^th^ sample measures 2.0-fold higher, and the 7^th^ sample 1.6-fold higher, that is, both were overexpressed by more than 1.5-fold (see Table 1A). The 8^th^ and 9^th^ samples were obtained from breast cancer patients, one from a urine sample, and the other from a serum sample with let-7b copy numbers 1.77 HL and 1.80 HL. Both measurements were 2.0-fold higher than those of the control 0.89 HL. These data suggest that 1.5 may serve as a threshold, whereby an miRNA level above 1.5 HL suggests cancer detection, and an miRNA level below 1.5 HL indicates the absence of cancer. Additional experiments were conducted with additional miRNAs, where a 1.5-fold threshold was applicable.

The number of **miR-15b copies** in the serum of H6914 and in the sera of healthy individuals and cancer patients was found to be directly proportional to the RNA content in the range of 9.1 to 20.7 ng/μL [37]. This observation was reported earlier ([37] 8 samples, four healthy and four diseased), confirmed here, and extended by including serum and urine data (13 new samples, eight healthy and five diseased) in the range of 6.9 to 174.6 ng/μL RNA (Table 4). These data suggest that miR-15b is not a cancer biomarker, in agreement with previous findings [20]. The observed independence in age, sex, or ethnicity is in bold contrast to studies that report large data variation and attribute it to age, sex, ethnicity, and other parameters. Our data irrevocably established that miRNA copies must be normalized to the same RNA content, which is currently not common practice.

**Normalization to the same RNA content** (16.0ng/μL in H6914, 1^st^ lot) for all tested miRNAs yielded copy numbers independent of age, sex, ethnicity, and cancer indication (breast, prostate, or pancreas), as well as biospecimen, suggesting that a urine sample may replace a blood draw. Further normalization, that is, dividing the miRNA copies of a sample by the corresponding miRNA copies from H6914 (1^st^ lot), yielded two groups with zero overlap, one averaging 1.01 HL with RSD=0.16 (40 counts: all miRNAs from healthy samples + miR-15b from cancer samples) and another averaging 1.83 HL with RSD=0.09 (28 counts: miR-21, miR-375, and miR-141 from cancer samples) (Table 4 and Figure 2). On the first inspection, the data yielded 100% sensitivity and 100% specificity; however, the sample size was small, and whether healthy subjects would remain free of cancer and for how long was not assessed. Notably, a p-value of 1.6×10^-22^ was determined by Excel’s t-test for the combined three cancer biomarkers in the healthy vs. the cancer group (sample size 52). For comparison, p-values of approximately 0.001 were used for miR-141 measurements by ddPCR, which is currently considered the most accurate method [27,32]. Despite the small study size, the unprecedented discrimination observed between healthy samples and samples from patients with breast, prostate and pancreatic cancer validates each of these three miRNAs as biomarkers for cancer. Overexpression of miR-21 [25] and miR-141 [20] has been associated with numerous cancer indications, in addition to breast, prostate and pancreatic cancers. Further testing of this set of miRNAs in samples from additional cancer indications should illustrate their usefulness as multi-cancer biomarkers. As long as a biomarker, such as miRNA, is elevated by 80% or more between diseased and healthy samples, our protocol-defined ± 20% accuracy in miRNA copy number determination is compatible with a 1.5 HL threshold to assign an unknown sample as healthy or cancerous (Table 1A and Figure 2). The data in Table 4, in conjunction with miRNA studies conducted worldwide during the last 25 years, suggest that elevated levels of miR-21, miR-375, and/or miR-141 in the serum or urine warrant consultation with a doctor, like high fever would.

**Figure 2:**
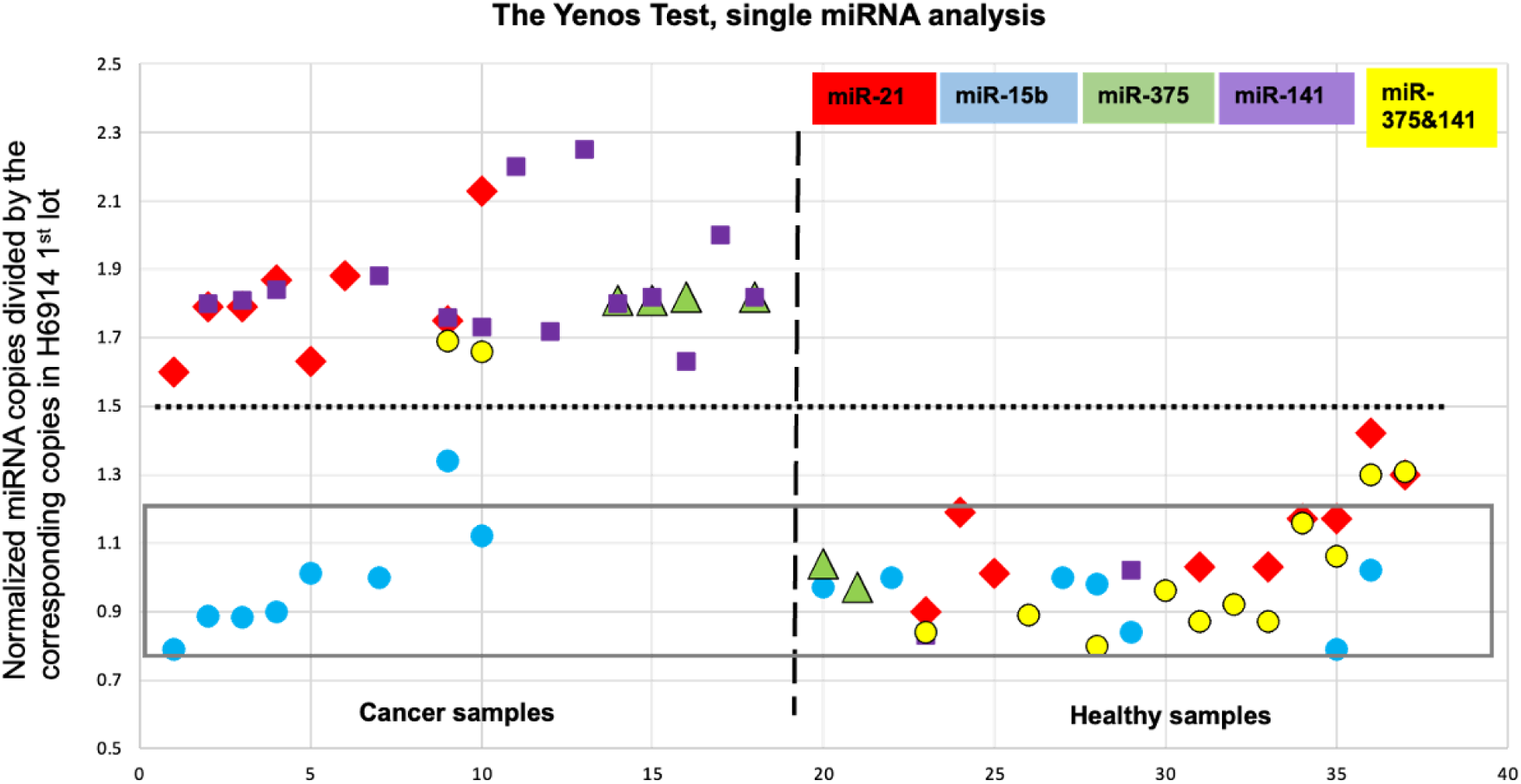
Data from Table 4, miRNAs per individual. miRNA copies were normalized to 16ng/μL RNA content and then divided by the corresponding miRNA copy number in H6914 1^st^ lot. This double normalization yields Level 1.00 for all 4 miRNAs measured in H6914 1^st^ lot (data not included in Tables or figure). The rectangle across samples with y-axis ranging from 0.8 to 1.2 (average HL = 1.00 and RSD = 0.2) includes 87% of the healthy data. The vertical dashed line separates cancer samples from healthy samples, whereas the horizontal dotted line at 1.5 HL is the threshold which discriminates healthy samples from samples with elevated levels of miR-21, miR-375 and miR-141 with a p value at of 1.6×10^-22^ (see discussion). The data confirms that the test exhibits no data overlap, i.e., sensitivity, specificity, PPV and NPV at 1.0 for each miRNA alone and for the miR-375 & miR-141 pair. Most importantly, this set of data tentatively validates each miRNA (miR-21, miR-375 and miR-141) as a cancer biomarker for all three breast, prostate, and pancreatic cancers. Additional samples are required to confirm validation.

**The nanopore technology used here** for trace nucleic acid detection and quantification uses a bracketing approach for measurement, making it unique; it has no similarities to the currently used assays. The assay was developed and optimized earlier [36,37,46]. To the best of our knowledge, no other analytical assay yields measurements with protocol-defined accuracy of ± 20%. When each measurement was 20% or better, a series of comparable measurements will exhibit RSD < 0.2. Each process involved in this assay is described in the Experimental Section and is outlined here. Total RNA (0.1 mL) was isolated from the serum (0.2 mL) using a Monarch kit from NEB, and total RNA (0.05 mL) was isolated from the urine (5 mL) using a Norgen slurry kit. The total RNA (ng/μL) was measured using a DS-11 DeNovix spectrophotometer. For accuracy reasons, isolated total RNA should contain more than 7 ng/μL RNA and have an absorbance ratio at 260 nm vs. 280 nm, A260/A280, better than 1.6. Capillary electrophoresis (CE) analysis, as described earlier [37], confirmed the dramatic variation observed in the total RNA isolated from individuals.

Samples for nanopore experiments were prepared by mixing a few μL of the isolated total RNA with a few μL of the probe complementary to the target miRNA. The mixture was stored at -20 °C overnight to ensure complete hybridization, and 75 μL of filtered ONT buffer was added to this mixture immediately before loading onto the MinION flow cell. The nanopore experiment was conducted for 45 min at -180 mV. The flow cell was allowed to rest for 15 min and a second run was conducted under the same conditions. MINKNOW software runs the experiments and produces a fast5 file (the ion current (i) with time (t)), which was subsequently analyzed using *OsBp-detect* [51]. The latter yielded a tsv file that was opened in Excel. The data were grouped in the form of a histogram with 0.05 bins (see Figures 3 and 4 in Materials and Methods). An experiment using a buffer only instead of a sample primed the flow cell and served as a control for the experiment with the RNA/probe sample. If the test with the mixture of RNA and probe is determined to be silencing, then the next test may be designed using the same aliquot of probe but only 67% of the RNA aliquot. If the second test was determined to be a detection experiment, then miRNA copies per μL of RNA sample were determined from the average of the probe copies per μL of RNA sample from the two test samples. Typically, more than two tests are necessary before finding the set, one detection and the other silencing, which fulfills the accuracy requirement. Owing to the low throughput of the assay and the approximately 15-hour life span of the flow cells, not all five miRNAs were measured in every sample. This platform may be further optimized by developing a buffer that is optimal for ion-conductance experiments to replace the currently used buffer. Buffer optimization may enable direct quantitation of miRNA targets from a single experiment by suppressing and/or controlling background noise.

**Figure 3:**
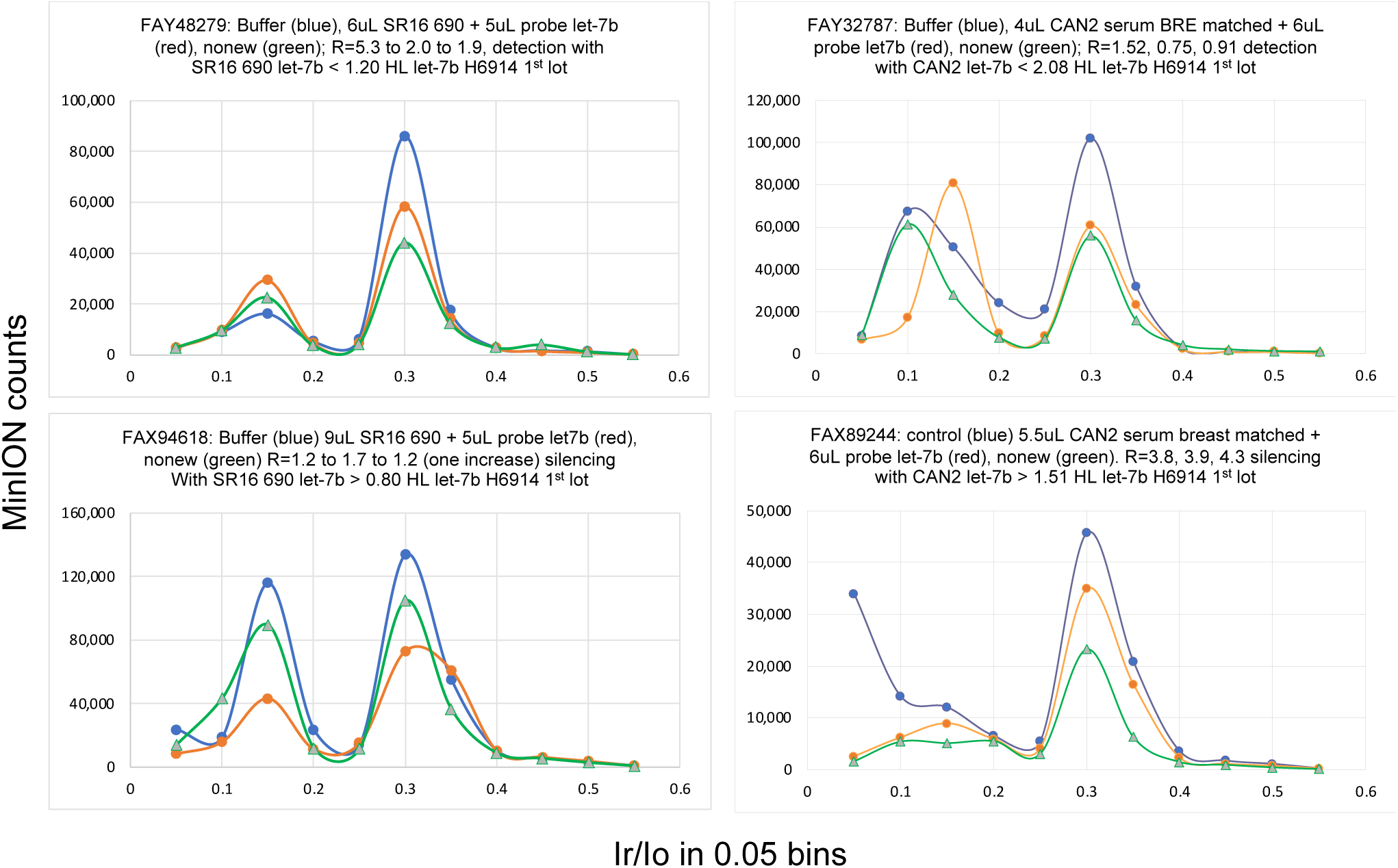
Examples of Yenos tests targeting let-7b taken from Table 3. Top figures illustrate detection experiments and bottom figures illustrate silencing experiments (see Data Analysis in Methods). Each figure shows a test, that is, a set of three experiments with buffer (blue), followed by the 1^st^ run of the sample which is a mixture of RNA with the probe (red), followed by a 2^nd^ run of the same sample. All three experiments were conducted at -180mV for 45 min. Analysis of the events by *OsBp_detect* typically yielded two maxima: one early Ir/Io=0.15 and a late Ir/Io=0.3. As shown, the buffer alone exhibited events at both maxima, but the Yenos probes traversed only at Ir/Io=0.15. Therefore, the presence of a free probe is consistent with an increase in the early Ir/Io peak and/or a decrease in the late Ir/Io peak because there is a steady decrease in events due to the inactivation of the nanopores. Silencing experiments (bottom) often exhibit a markedly reduced number of events due to nanopore “shielding” as discussed above, while detection experiments (top) exhibit comparable counts but a reversed distribution with relatively more events at the early *(I_r_/I_o_)_max_* = 0.15 and fewer events at the late *(I_r_/I_o_)_max_* = 0.30. For a specific experiment with an x μL probe and y μL of sample RNA, probe molecules P = x μL (probe concentration in fM) × 600. If the experiment involved detection, then P > target miRNA molecules within the y μL aliquot. If the experiment involved silencing, then P < target miRNA molecules in y μL. It follows that the number of miRNA molecules per 1 μL of isolated RNA sample < or > P/y, depending on the experimental outcome.

**Figure 4:**
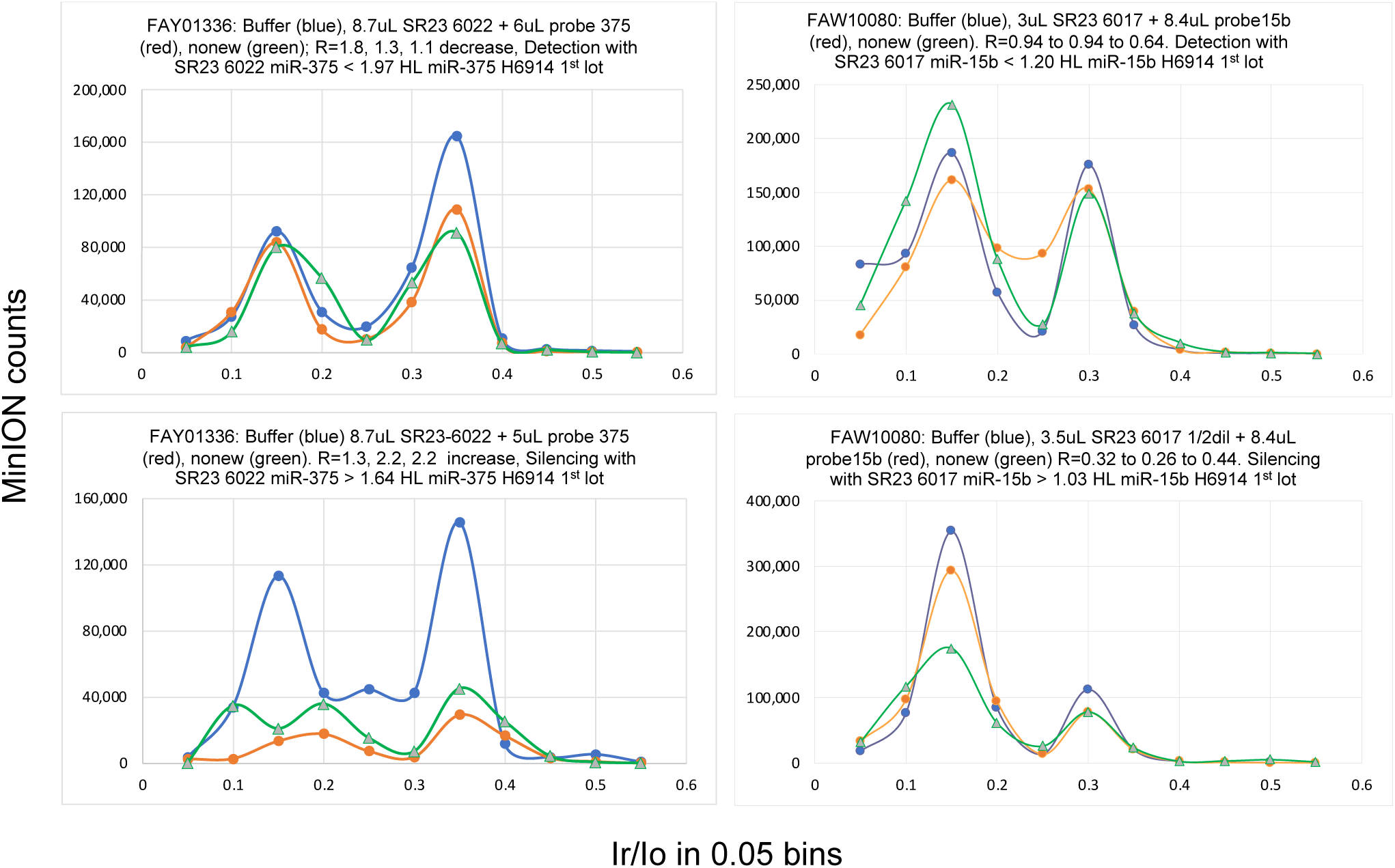
Examples of Yenos tests targeting miR-375 and miR-21 taken from Table 4. Top figures illustrate detection experiments and bottom figures illustrate silencing experiments. For additional information see Data Analysis in Methods and the caption of Figure 3.

**A solid-state nanopore array** is a more robust and cost-efficient alternative to the proteinic nanopores. Our technology can be implemented in other nanopore arrays with no or minor optimization, as shown using α-hemolysin [45] and silicon nitride nanopores [44]. The only requirement for this assay is that the width of the nanopore permits the translocation of ss nucleic acids and prevents the translocation of ds nucleic acids. Interestingly, the “bulkier” probe traverses nanopores of the same width as the unlabeled (intact) nucleic acids. We attribute this observation to the osmium tags extending parallel to the strand axis and not perpendicular to it. This configuration reduces the number of water molecules carried by the nucleic acids. For thermodynamic reasons, a naked ss nucleic acid (with no water solvation) is unlikely to exist in water, and applying a voltage cannot eliminate the 1^st^ solvation/hydration shell. The decreased hydration of the probe is envisioned as decreased “lubrication” during the sliding of the probe through the nanopore wall and may rationalize the dramatically slower translocation.

### This technology is not limited to the five miRNAs measured here

The probe is a DNA oligo complementary to the target sequence. Osmium tagging is straightforward, and the resulting probe is stable and characterizable (see Methods section). The design of the probe is general and has been optimized for efficient hybridization with a DNA or RNA target, and for nanopore detection. Probes for a limited number of human miRNAs have been manufactured and tested (Table 6 and [37]), and miRNAs from other species can also be targeted. However, this technology is not limited to miRNA quantification. Circulating tumor DNA (ctDNA), circular nucleic acids, and practically any ss nucleic acid of a known partial sequence of interest can be detected and quantified. Liquid biopsies are non-invasive; however, this assay is not limited to liquid biopsies. Because of the availability of commercial kits for total RNA isolation (including miRNAs) from practically any tissue or organ, the latter can be used for ss nucleic acid detection and quantification.

**Table 6:**
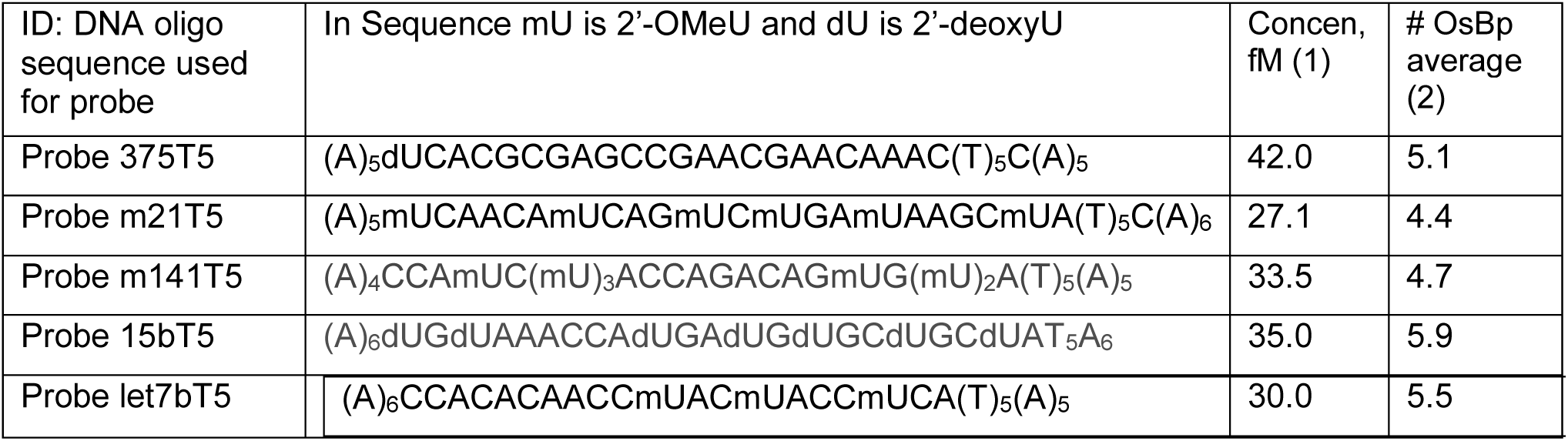
Sequence and characterization of the probes used in this work. (1) Concentration of probe solution (fM) used for the nanopore experiments. It was obtained by 5/1000 or 10/1000 dilutions from the stock solution (μM) of the probe prepared by osmylation (T-OsBp)_5_ of the oligo and characterized in-house by HPLC. (2) The average number of osmium label moieties on the probe (extent of osmylation) was measured using the following equation: absorbance at 312 nm/absorbance at 272 nm or R(312/272) = 2x(no osmylated pyrimidines/total nt) [37]. R is the ratio of the corresponding HPLC peaks, regardless of their shape (sharp or broad). An extra osmium tag was conjugated to a C or U base within the sequence. A single internal tag did not prevent hybridization, as shown by nanopore experiments.

**Implementation of a multi-cancer test** will not require extensive testing, such described in Table 3, which is suitable for miRNA validation studies. Instead of determining miRNA copy number, the concept of threshold value may be implemented as follows: considering an miRNA biomarker, like the ones studies here where the HL level is close to 1.0, and the cancer level is close to 1.8, both at ∼0.2 RSD, an experiment designed to target a miRNA level at ∼1.5 HL threshold (Yenos test) should yield detection with healthy samples and silencing with cancer samples or samples from asymptomatic individuals with elevated miRNA (Tables 1A, 3, 4 and Figure 2). We are proposing that elevated miRNA levels in two out of three tested miRNA cancer biomarkers, indicate miRNA dysregulation which may be associated with the onset and/or presence of cancer and represent a warning sign for the tested individual. The number of false-positive and false-negative results from such a multi-cancer test targeting ∼1.5 HL should be practically nonexistent, as shown in Tables 1A, 3, and 4 and Figure 2. A single miRNA test included a control/baseline experiment and two runs using the same sample (see Methods). A second test for an additional miRNA will include one control and a second sample run twice. Six separate experiments will be performed for two miRNA-related cancer biomarkers at the ∼1.5 HL threshold, and the results should collectively lead to one conclusion, namely, whether the biomarkers were detected or silenced (Figures 3 and 4). We exploited this approach by testing consented individuals and showed that it works. One consented individual was a breast cancer survivor and her miRNA levels, tested a few years later, were comparable to those of the healthy controls, suggesting that miRNA levels recovered. Another individual who tested positive for cancer in the Yenos urine test, underwent the Galleri test, which also exhibited cancer detection, thus confirming the Yenos test. Notably the Galleri test evaluates DNA methylation, whereas the Yenos test evaluates miRNA overexpression, making them scientifically and technically independent. All four miRNAs tested exhibited comparable overexpression in the cancer samples (1.5 HL). Other miRNAs may exhibit a comparable or different overexpression and if overexpression is at least 1.8-fold, our technology with a protocol-defined RSD ∼0.2 will discriminate healthy from diseased samples.

**Table 3 lists the results of the experiments in which two miRNAs, miR-375 and miR-141** were simultaneously targeted using the two corresponding probes. Targeting two miRNAs in one experiment yielded one copy number for both miRNAs, which reduced the number of experiments two-fold and could still serve as a screening test using an appropriate threshold value, as outlined above. Figure 2 illustrates the successful use of combined miRNAs to discriminate between cancerous and healthy samples. This approach can only be used when two targeted miRNAs exhibit similar copy numbers, as is the case for miR-375 and miR-141. This limitation is due to the inability of MinION nanopores to discriminate one probe from another with the current probe design (Table 6). Earlier work with α-hemolysin [45] and solid-state nanopores [44] illustrated that more osmium tags yielded deeper translocations with an earlier (*I_r_/I_o_*)_max_. Adding another 2-3 osmium tags to the current five tags may yield the desired discrimination and enable individual miRNA testing in a single test targeting two miRNAs

## CONCLUSIONS

This report presents the implementation of a novel analytical platform for the detection, quantification, and validation of miRNA cancer/disease biomarkers from liquid biopsies with an accuracy of ± 20%. This technology combines single-molecule ion conductance experiments using MinION functionality with an expertly optimized probe design. A general validation strategy applicable to any potential ss nucleic acid biomarker is presented. The copy numbers of five miRNAs, let-7b, miR-15b, miR-21, miR-375 and miR-141, were measured in the healthy and cancerous samples. Normalization of the copy number to the same RNA content was found to be critically important. miRNA copies from the combined serum of healthy men (H6914) were found to be equivalent to the miRNA copies measured in urine samples of healthy subjects, men, and women, clearly illustrating the equivalence between serum and urine for the tested miRNAs. In contrast to miR-15b, which appears to be unrelated to breast, prostate and pancreatic cancers, and let-7b, which is not overexpressed in pancreatic cancer, the other three miRNAs were elevated in all three cancers, in agreement with the findings of multiple studies conducted over the last 25 years. The 1.8-fold overexpression of these cancer biomarkers agrees well with the overexpression observed earlier in a prostate cancer study [23]. In addition, the 1.8-fold overexpression of miR-21 is in excellent agreement with the 1.7 overexpression observed in a lung cancer study [22]. Strikingly, the normalized copy numbers of each of the five miRNAs appear to be independent of age, sex, and ethnicity, in bold contrast to the variability observed using current miRNA quantification platforms. Compared with the corresponding miRNA copy number from the combined serum of healthy men (H6914 from Sigma‒Aldrich), the data were grouped into healthy and cancer samples with no data overlap, that is, zero false negatives and zero false positives, yielding sensitivity, specificity, positive predictive value (PPV) and negative predictive value (NPV), all equal to 1. This unprecedented discrimination tentatively validated each miRNA (miR-21, miR-375 and miR-141) separately, as three-cancer biomarkers. The technology merits further testing in a larger sample size, as well as for other indications in addition to breast, prostate, and pancreatic cancers. The ability of this platform to accurately quantify ss nucleic acid traces, to validate potential miRNA biomarkers, and its prospective adaptability to future solid-state nanopore platforms is unprecedented.

## MATERIALS AND METHODS

### Human samples

Human serum from the USA, isolated via sterile filtration from male AB-clotted whole blood (H6914, 1^st^ lot SLCH8785, 2^nd^ lot SLCJ3635; data reported earlier; 3^rd^ lot SLCL6534 and 4^th^ lot SLCN9213; data reported here in Table 2) were purchased from Sigma‒Aldrich over a period of 3 years. Serum samples purchased from Discovery Life Sciences (DLS, Huntsville, AL, USA) and Tissue for Research (Accio Biobank online, Suffolk, UK) were collected from informed consented individuals under the IRB/EC protocol. The selection of these samples from a large depository included both male and female donors, if applicable, and one each from African American, Hispanic, or White ethnicity. Samples were collected from newly diagnosed, naïve, and pre-treated patients. The demographic information of the patients with cancer who provided their specimens is listed in Table 5. The project to include Urine samples collected from consenting healthy subjects was reviewed by the Advarra Investigational Review Board (IRB). The protocol and consent form were reviewed, modified, and approved by the Advarra IRB on November 15, 2023. Protocol: Yenos Analytical LLC-02. Quantification of selected microRNAs in the urine of healthy individuals (Pro00074065). Donors of urine samples reviewed and signed an informed consent form. They were then sent a kit/insulated box, cold bricks, and instructions to collect their biospecimen at home, freeze it and ship it overnight to the Yenos facilities. The healthy urine donors were 30 to 75 years old, female, or male and of different ethnicities. For the isolation of total RNA from serum, we used the Monarch T2010S Kit (1^st^ lot 10075450, 2^nd^ lot 10141109 reported earlier, and 3^rd^ lot 10144556 used here). For the isolation of total RNA from urine, two kits were used: No. 29000 was used for 1 mL urine samples, and the slurry kit No. 29600 was used for 5 to 10 mL urine samples. All kits were used according to the manufacturer’s instructions.

### Oligos, Probes, and other Reagents

The only ONT kit used for the experiments reported here was the Flow Cell Priming Kit XL (EXP-FLP002-XL), ONT flush buffer or ONT buffer. The ONT buffer is proprietary, provides the necessary electrolytes and must represent more than 80% of the approximate 80 μL sample volume. Custom-made DNA oligos and 2’-OMe-oligos synthesized at the 0.2 μM scale and purified by HPLC or PAGE by the manufacturer were purchased from Integrated DNA Technologies (IDT) and Millipore/Sigma‒Aldrich, respectively. Oligos (sequences in Table 6) were diluted with Ambion nuclease-free water, untreated with DEPC, typically to 100 or 200 μM stock solutions, and stored at −20°C. The oligo purity was confirmed to be >85% by in-house HPLC analysis [55]. Following osmium tagging (osmylation, see below), in-house HPLC analysis was used to determine the probe content, extent of osmylation, and efficiency of probe/target hybridization [37]. Osmylated oligos were manufactured in-house, based on published methods at a concentration of approximately 30 μM. LoBind Eppendorf test tubes (1.5 mL) were used for serial 5/1000 or 10/1000 dilutions to yield probes at concentrations of 15-30 fM. Mixtures of probe with isolated RNA were prepared in 0.5 mL RNase-, DNase-free, sterile test tubes and stored at -20 °C overnight.

**Osmylation of nucleic acids** using a 1:1 mixture of OsO_4_ and 2,2’-bipyridine, abbreviated OsBp, was discovered 60 years ago [56], used extensively [57–59], and optimized by us [60,61]. The detailed protocols for the synthesis, purification and quality control assays have been previously described [36,37]. OsO_4_ is a hazardous material, and care must be taken for its use, storage, and disposal [62]. Osmylation reactions require a 20-fold excess of OsBp over the reactive pyrimidine in monomer equivalents to ensure pseudo-first-order kinetics and to yield preferential labeling of thymidines (T) over the other pyrimidines. The osmylation reaction was quenched upon purification. Purification from excess OsBp (twice) was performed using spin columns (TC-100 FC from TrimGen Corporation) for 4 min at 5,000 rpm. The flow-through solution is a probe that is chemically stable that can be stored at -20°C for two years.

**The development, optimization, and validation** of probes for enhanced MinION detection have been previously reported [37]. The optimized probe comprises a sequence complementary to its target but extended at one end with four to five adjacent T residues and flanked by up to five adenosines (A) at either end (Table 6). This facilitated the entry of the probe into the nanopores. The adjacent Ts were tagged with an osmium label for quantitative detection. Within the probe sequence complementary to the target, Ts is replaced by uridine (U), 2’-OMe-U or dU to minimize OsBp labeling because the osmylation kinetics of U and cytosine (C) are substantially slower than that of T [60]. HPLC analysis yields the probe concentration (content) using intact oligo as a standard because the absorbance of the probe at 260 nm is practically the same as that of the precursor intact oligo [60]. HPLC analysis provided evidence of the quantitative depletion of the OsBp reagent. Alternatively, a suitable spectrophotometer can be used to determine the content and extent of osmylation.

### Single-molecule ion-channel conductance experiments on the MinION (MinION Mk1B platform)

One must register with the ONT and download the software MinKNOW to a computer/laptop with specifications provided by ONT. All the functions necessary to test the hardware and flow cells and run the experiments were performed using the MinKNOW software. The sample was loaded onto a flow cell that fitted within the MinION device. The experiment was run under “start sequencing” mode. A direct RNA sequencing kit (SQK-RNA002) was used to initiate experiments. The flow cell type FLO-MIN106 was selected, and the run length (45 min) and bias voltage (-180 mV) were selected; basecalling was disabled, and the output bulk file Raw (1-512) was checked and generated. The output location was Library/MinKNOW/data/, and the output format was *fast-5*. All the experiments reported here were run for 45 min at -180 mV. The *fast-5* file was analyzed using the *OsBp_detect* algorithm [51]. The number of events per channel from the *OsBp_detect* analysis was compared with the actual *i-t* trace of the specific channel using MATLAB visualization, and this algorithm, 2^nd^ revision, was repeatedly validated. Currently *OsBp_detect* can only be used with a 2017 or an earlier version of MacBook Pro loaded with macOS 10.14 Mojave. Future work will include the adaptation of *OsBp_detect* to newer operating systems. While alternative parameters were explored, all experiments reported here were analyzed using the following threshold parameters: (i) event duration (in tps): 4-1200 (1.3-400 ms), (ii) lowest *I_r_/I_o_*<0.55, and (iii) all *I_r_/I_o_* <0.6, channels 1-512.

### Data analysis

A state-of-the-art laptop/computer requires approximately 5 min for *OsBp_detect* analysis, produces a file in *tsv* format, opens via Microsoft Excel and saves it as such. In the Excel spreadsheet, the algorithm-selected events (*I_r_/I_o_* data) are grouped in the form of a histogram with 0.05 bins, from 0.05 to 0.55, and plotted (Figures 4 and 5). These events were added together and identified as *Total Events*. Typical histograms exhibit two maxima *(I_r_/I_o_)_max_*: an early one at *I_r_/I_o_* =0.15 and a late one at *I_r_/I_o_*=0.30. These maxima may vary by ±0.05 units depending on the flow cell age. The events under late *(I_r_/I_o_)_max_* and early *(I_r_/I_o_)_max_* were noted, and their *ratio ((I_r_/I_o_)_max_ late (0.3)/early (0.15))* was calculated. These values (*total events and ratio (R)*) represent the criteria by which an experiment is judged as detection or silencing compared to the buffer control. Fewer total events than those in the control suggested silencing, whereas more total events suggested detection. A decreasing *ratio* R indicates detection, and an increasing *ratio* R indicates silencing. This assignment is consistent with an increased number of events owing to the presence of the probe, which traverses with *(I_r_/I_o_)_max_* ∼ 0.15, whereas intact RNA and background noise traverse mostly with *(I_r_/I_o_)_max_* ∼ 0.35. Each sample was run twice, and each run was compared to the buffer/control. Because the flow cells lost active pores during every experiment, the *total* number of events decreased during every experiment. Fewer nanopores reduced the effect of events owing to the free probe. In contrast, fewer nanopores enhance the effect of fewer events, owing to the absence of the probe. The presence of the hybrid had an additional effect on reducing the number of events. This is because the hybrids are driven toward the nanopores, cannot pass through, and are pushed back by the alternating voltage of the platform. However, they remain in proximity and prevent other molecules from traversing the nanopores.

## Data availability

The data generated during this study are included in this published article and/or in the Supplementary Section. Raw data (*FAST-5* format at ∼3.3 GB each) may be obtained from AK.

## Acknowledgments

This study was supported by private funding. We are grateful to Dr. Albert Kang for developing the *OsBp-detect* algorithm specifically for this application. We thank Professor Miten Jain from Northeastern University for sharing the algorithm used to visualize the *i-t* trace of each channel of the *fast-5* file using MATLAB from MathWorks.

## Competing Interests

Anastassia Kanavarioti is the Founder and Director of Yenos Analytical LLC, a company that delivers custom analytical solutions for native, synthetic, or transcribed nucleic acids and engages in the development and manufacturing of labeled/tagged nucleic acids for use in conjunction with nanopore detection and nucleic acid quantification, including miRNAs, with awarded patents for *Nanopore platform for DNA/RNA oligo detection using an osmium tagged complementary probe,* Patent No: US 11,111,527 B1, issued 9-7-2021 and US 11,427,859, issued 9-30-2022 and *Osmium Tagged Probes for Nucleic Acid Detection,* US 11,884,968, issued 01-30-2024. In 2023, international patents were filed in Europe, Canada, Australia, Japan, China, India, Brazil, and South Korea. A provisional patent application entitled *Detection, Quantification and Validation of microRNA Biomarkers* was filed with USPTO on May 7, 2024. There are no competing interests or relationships (commercial, financial, or nonfinancial) between the two companies, namely Oxford Nanopore Technologies and Yenos Analytical LLC, and their employees.

## Notes

### Funding Statement

This study did not receive any funding

### Author Declarations

Ethics committee IRB Advarra waived ethical approval for this work. The project to include Urine samples collected from consenting healthy subjects was reviewed by the Advarra Investigational Review Board (IRB). The protocol and consent form were reviewed, modified, and approved by the Advarra IRB on November 15, 2023. Protocol: Yenos Analytical LLC-02. Quantification of selected microRNAs in the urine of healthy individuals (Pro00074065). Donors of urine samples reviewed and signed an informed consent form. They were then sent a kit/insulated box, cold bricks, and instructions to collect their biospecimen at home, freeze it and ship it overnight to the Yenos facilities.

